# Germline mutations in hereditary breast cancer genes are associated with early age at diagnosis and family history in Guatemalan breast cancer

**DOI:** 10.1101/2020.08.09.20171165

**Authors:** Megan Ren, Anali Orozco, Kang Shao, Anaseidy Albanez, Jeremy Ortiz, Boyang Cao, Lusheng Wang, Lilian Barreda, Christian S. Alvarez, Lisa Garland, Dongjing Wu, Charles Chung, Jiahui Wang, Megan Frone, Sergio Ralon, Victor Argueta, Roberto Orozco, Eduardo Gharzouzi, Michael Dean

**Affiliations:** Division of Cancer Epidemiology and Genetics, National Cancer Institute, NIH, Gaithersburg, MD USA; Instituto Cancerologia, Guatemala City, Guatemala; BGI-Shenzhen, Beishan Industrial Zone, Shenzhen 518083, P.R. China; Department of Computer Science, City University of Hong Kong, Kowloon, Hong Kong SAR, P. R. China; Hospital General San Juan de Dios, Guatemala City, Guatemala; Cancer Genetics Research Laboratory, Division of Cancer Epidemiology and Genetics, Frederick National Laboratory for Cancer Research, Gaithersburg, MD USA; Integra Cancer Center, Guatemala City, Guatemala

## Abstract

**Background:** Mutations in hereditary breast cancer genes play an important role in the risk for cancer, however, little is known of the type and frequency of mutations in Central American populations, including Guatemala.

**Methods:** Two separate panels of known cancer susceptibility genes were used to sequence blood DNA from 664 unselected breast cancer cases from two large hospitals in Guatemala. Variants were annotated with ClinVar and VarSome. Data from a structured questionnaire was used to compare mutation carriers of medium and high penetrance genes.

**Results:** A total of 73 out of 664 subjects (11%) had a variant classified as pathogenic in a gene with known high or medium penetrance for inherited breast cancer. The most frequently mutated genes were *BRCA1* (37/664, 5.6%) followed by *BRCA2* (15/664, 2.3%), *PALB2* (5/664, 0.8%) and *TP53* (5/664, 0.8%). Pathogenic variants were also detected in the moderate penetrance genes *ATM*, *BARD1*, *CHEK2*, and *MSH6*, and rare pathogenic variants detected in the low penetrance genes *AXIN2*, *FH*, *MLH1, MSH2, MUTYH*, *NF1*, and *SDHB*. The high ratio of *BRCA1*/*BRCA2* mutations is due to the presence of two potential founder mutations, *BRCA1 c.212+1G>A* splice mutation (15 cases) and *BRCA1 c.799delT* (9 cases). Compared to all others, cases with pathogenic mutations had a significantly earlier age at diagnosis (45 vs 51 years, P<0.001), more likely to have had diagnosis before menopause, and a higher percentage had a relative with any cancer (51% vs 37%, P=0.038) or breast cancer (33% vs 15%, P<0.001). Mammography usage was less frequent in lower SES women indicating this group is less likely to be screened for breast cancer (p < 0.001).

**Conclusions:** Guatemalan women have rates of hereditary breast cancer mutations similar to other populations, and these women are more likely to have early age at diagnosis and family history. This data supports the use of genetic testing in breast cancer patients and those at high risk as part of a strategy to reduce breast cancer mortality in Guatemala.

## Introduction

Breast cancer is the most commonly diagnosed cancer among women worldwide. In Latin America and the Caribbean, female breast cancer accounts for 15% of all cancer cases among women in 2018 [1]. Although incidence rates of breast cancer are higher in high income countries, low-and-middle-income countries (LMIC) experience higher mortality rates partially explained by limited healthcare access and late stage diagnosis.

Genetic testing for germline mutations in breast cancer susceptibility genes can identify individuals with a higher risk of developing breast cancer, but there is limited information on the mutation profile of many Latin American populations [2, 3] Hall et al. documented that Latin American women referred for genetic testing have equal or higher rates of *BRCA1* and *BRCA2* mutations (15%) as other groups [4]. The frequency and spectrum of hereditary cancer gene mutations have been described in Latin American and US Hispanic populations [5-8]. However, cancer disparities among US Hispanic subgroups may be further masked because they tend to be aggregated as a monolithic group in epidemiological studies.

With a population of approximately 18 million people, Guatemala is the most populated country in Central America. The demographics in Guatemala are reflective of its indigenous and Spanish influence. According to data from the World Population Review, 41% of the population in Guatemala has mixed European and indigenous ancestry, 39% are full Amerindian, and 19% are whites of European descent [9]. Guatemala has a higher percentage of Amerindian individuals than any other nation in the Western hemisphere and most indigenous people are Mayan [9]. Those with European descent can generally trace their ancestry back to early Spanish and German settlers.

Currently, Guatemala has one of the poorest health indicators in Latin America, with a high infant mortality rates (~28 deaths per 1,000 live births in 2010-2015) and low life expectancy (~75 years), both of which are directly associated with unequal wealth distributions [10, 11]. Inequality and poverty in Guatemala remain consistently high, with approximately 50% of the population falling under the poverty line and 9% of those are affected by extreme poverty [12]. In regard to breast cancer, higher income and socioeconomic status are associated with increased breast cancer incidence and decreased mortality [13]. According to estimations from Globocan in 2018, breast cancer in Guatemala accounts for 19% of cancer cases among women, followed by cervical cancer at 16%, stomach cancer at 11.7%, and liver cancer at 10% [1]. In addition, breast cancer has the highest incidence rate of any cancer and it has the fourth highest mortality rate after stomach, liver, and cervical cancers among Guatemalan women [1]. Among US Hispanic females, breast cancer is the most common cancer for both estimated new cases and cancer-related deaths at 29% and 16% respectively [14].

The *BRCA1* and *BRCA2* genes are highly penetrant susceptibility genes for hereditary breast cancer. Inactivating mutations in these genes occur in approximately 5-10% of all breast cancers [15]. Recurrent mutations, or mutations identified in more than one unrelated BRCA carrier, have been described such as *BRCA1* 185delAG, *BRCA1* 5382insC, and *BRCA2* 6147delT that explain 78% of cases in Ashkenazi Jews [16]. In Latin America, some variants such as *BRCA1* 185delAG and R1443X are among the 20 most frequent *BRCA1* variants reported by the Breast Cancer Information Core database, while others like *BRCA1* A1708E are among the 10 most frequent pathogenic variants in Latin America but not overall [3]. This study analyzes the breast cancer mutation profile of women from Guatemala where few genomic studies for breast cancer currently exist. By increasing scientific knowledge of breast cancer susceptibility genes in Guatemalan women, the results from this study can inform clinical diagnostics for Guatemalan women both abroad and in the U.S.

## METHODS

### Study design and data collection

This study was conducted at the Hospital General San Juan de Dios, a large public hospital and the Instituto de Cancerología (INCAN) an adult cancer hospital in Guatemala City. The Research Ethical Committees of each institution approved the protocol, and the study was determined exempt from institutional review board (IRB) approval by the NIH Office of Human Studies Research. Women attending either of these hospitals for their breast cancer diagnostic biopsies were invited to participate and gave written informed consent. Two 5 ml tubes of blood were collected and frozen at -20°C as well as a tumor biopsy stored in 0.5 ml of RNAlater solution at -20oC. Trained interviewers administered an approved questionnaire including reproductive history, family history of cancer, and socioeconomic data. Mammography use was calculated considering only patients over the age of 40, we determined the difference between the age of diagnosis and age at a first mammogram. If the difference was two or greater, we assumed the patient received regular mammogram exams and had one within the past two years. To estimate socioeconomic status (SES) the lack of job security makes it difficult for subjects to estimate their annual income; thus, we took cookstove type to be an indicator of SES due to the known association between wood-burning stoves and poverty in Guatemala, particularly among indigenous Mayans [17].

### Sample preparation and whole-exome sequencing

DNA was extracted from the participant’s peripheral blood samples by a Qiagen DNA Blood Mini kit (Qiagen, Hilden, Germany) according to the manufacturer’s instructions. Qubit Fluorometer (Life Technologies) and agarose gel electrophoresis were used to detect DNA concentration and integrity.

### Next-Generation Sequencing and Variant Calling

Gene target regions were captured, and DNA sequencing was performed at the National Cancer Institute and BGI Shenzhen. The samples were separated into two groups and analyzed on different sequencing platforms. A total of 587 samples from the Instituto Nacional de Cancerología (INCAN) were sequenced on the BGISEQ-500 platform (MGI, a BGI Company) with Paired-end 100 bp and 86 samples from Hospital General San Juan de Dios were sequenced on the Hiseq 2500 platform (Illumina) with the Paired-end 200 bp strategy. The genes on each panel are show in **Supplementary Table 1**.

For the samples on BGISEQ-500 platform, the DNA amount used for library construction was 1ug. DNA was randomly fragmented to 200-400bp by Covaris E210 (Covaris Inc.), and DNA libraries were constructed by end-repair, poly-A tailing, adapter ligation, and PCR amplification. The coding region and coding region ±30 boundaries of 115 genes were captured by a BGI capture array (produced by BGI). DNA libraries were subjected to BGISEQ-500 for sequencing with pair-end 100bp strategy according to the manufacturer’s protocol. The average depth of samples is over 650X (235x at least) and 99% coverage on target regions. Over 1.0G base of data was generated for each sample. Reads were filtered with SOAPnuke 1.5.0 and assembled with BWA 0.7.12. Bam file is processed with Samtools 1.2 and duplication was marking with MarkDuplicates 1.138 with standard parameters. GATK 3.4 was used to perform base quality recalibration and local realignment. Germline mutations were called with GATK 3.4 and filtered by quality depth, strand bias, mapping quality and reads position. (major parameters were -filter “QD < 2.0 || FS > 60.0 || MQ < 40.0 || SOR > 4.0 || MQRankSum < -12.5 || ReadPosRankSum < -8.0” for SNP_filter, and -filter “QD < 2.0 || FS > 200.0 || ReadPosRankSum < -20.0 || SOR > 10.0” for INDEL_filter).

For the samples on the Hiseq platform, genomic DNA with the initial amount above 1 ug was randomly fragmented to 200-300bp by Covaris E210 (Massachusetts, USA). Then the library was constructed as follows: end-repair, A-tailing, adapter ligation, and PCR amplification. PCR products were captured and quantified by quantitative PCR and pooled for sequencing on the Hiseq 2500 (Illumina) according to the manufacturer’s protocols (https://support.illumina.com/content/dam/illumina-support/documents/documentation/system_documentation/hiseq2500. Over 0.6G base data was generated for every sample with an average depth of about 200X and over 99% coverage on target regions.

Variants were annotated using automated pipelines and potential pathogenic variants identified. For our initial analysis, we included known breast cancer susceptibility genes *BRCA1*, *BRCA2*, *PTEN*, *TP53*, *PALB2*, *CDH1*, *ATM*, and *CHEK2*. Further validation was performed by manual review using the Integrative Genomics Viewer (IGV) [18]. Variants were classified as frameshift, stop-gain, splicing, in-frame deletion/insertion, or nonsynonymous and missense. The pathogenicity of each mutation was then analyzed using online databases including ClinVar and Varsome. Finally, the variants were compared with corresponding clinical data for population-wide trends.

### SNP Microarray and Ancestry Analysis

To determine the genetic ancestry of the samples 100ng of DNA was labeled and hybridized the Infinium OncoArray-500K BeadChip (Illumina) and data processed by the standard Illumina microarray data analysis workflow. Genotypes were analyzed with PLINK [19] to confirm gender (all samples were confirmed female), confirm three expected duplicates, and identified five unexpected duplicates (subjects recruited at both INCAN and HGSJDD). One of each unexpected duplicate was excluded.

The genetic ancestry of patients was estimated using a set of linkage disequilibrium pruned markers and running SNPWEIGHTS software with the reference panel provided containing the following populations: European, West African, and East Asian (ASN) [20]. The association between ASN ancestry was correlated with self-reported ancestry in a Guatemalan reference population (Supplementary Table 2). Ancestry was further confirmed using GRAF-POP [21].

### Statistical analysis

Median and interquartile ranges [IQRs] were calculated for the continuous variables, while frequencies and percentages were computed for the categorical variables. Wilcoxon rank-sum test was used to examine differences between age at diagnosis, age at menarche, age at first pregnancy, number of children, number of pregnancies, and number of abortions with presence of pathogenic mutations in SAS software v 9.4 (SAS Institute, Cary, NC). In addition, two proportion Z-test (two-tailed) was used to assess the difference in the percentage of patients having a family history of cancer, family history of breast cancer, contraception use, and NCCN status. A Chi-squared test with Yates correction was used to examine the relationship between socioeconomic status (cookstove type) and mammogram screen usage. In all calculations, a p-value of 0.05 or less was deemed significant.

## RESULTS

### Study Population

In total, 664 patients with histologically confirmed breast cancer were recruited from the Instituto de Cancerología and Hospital General San Juan de Dios, both in Guatemala City. Most of the patients self-identified as “Mestizo”, meaning non-indigenous people of mixed European-Amerindian ancestry who primarily speak Spanish [22]. Overall, the median age at diagnosis of the breast cancer cases was 49 (IQR: 41-61]) **(Table 1)**. The median number of children per study participant was 3 [IQR: 2-4] and the median age at menarche was 13 [IQR; 12-14]. Furthermore, fifty-seven percent of patients were post-menopausal at the time of diagnosis. Of the patients with available family history information, 17% had at least one first- or second-degree relative with breast cancer. The genetic ancestry of patients was determined using genotypes from a SNP microarray and compared to populations of European (EUR), Asian (ASN), or African (AFR) ancestry. Data from a separate population of Guatemalans, self-identified as indigenous, indicated ASN scores of 0.6 to be correlated with high indigenous ancestry **(Supplemental Table 1)**. In our study, ancestry data was available for 575 of the 664 patients. Of these 575 patients, 4.2% (24/575) had an ASN score of <0.2, 48% (273/575) had an ASN score between 0.2-0.4, 33% (191/575) between 0.4-0.6, and 15% (87/575) had an ASN score > 0.6. We chose to use ASN = 0.5 as a general cut off score for comparing patients with more or less indigenous ancestry.

**Table 1:** Association of study population characteristics with pathogenic mutations.

|  | Total, n = 664 |  | Pathogenic, n = 73 |  | Other, n = 591 |  | p-value |
| --- | --- | --- | --- | --- | --- | --- | --- |
|  | Median (IQR) | Range | Median (IQR) | Range | Median (IQR) | Range |  |
| **Age at diagnosis** | 49 (41-61) | 19-93 | 43 (37-52) | 23-86 | 50 (41-61) | 19-93 | <0.0001 |
| **Age at first pregnancy** | 21 (18-25) | 13-45 | 20 (17-25) | 15-35 | 21 (18-25) | 13-45 | 0.40 |
| **Age at menarche** | 13 (12-14) | 5-24 | 13 (12-14) | 9-18 | 13 (12-14) | 5-24 | 0.33 |
| **Number of children** | 3 (2-4) | 0-19 | 3 (2-4) | 0-10 | 3 (2-5) | 0-19 | 0.24 |
| **Number of pregnancies** | 3 (2-5) | 0-19 | 3 (2-4) | 0-10 | 3 (2-5) | 0-19 | 0.11 |
| **Number of miscarriages** | 0 (0-0) | 0-5 | 0 (0-0) | 0-1 | 0 (0-0) | 0-5 | 0.050 |

Population characteristics are compared between subjects with pathogenic mutations in High and Moderate penetrance genes and those with benign or no variants (Other). The median, interquartile range (IQR) are shown along with p-values for the comparison (Wilcoxon Two-Sample Test).

### Germline mutations in known breast cancer susceptibility genes

Targeted sequencing was performed on two different panels of 275 and 468 cancer susceptibility genes, including those known to cause inherited breast and ovarian cancer (*BRCA1, BRCA2, PALB2, PTEN, TP53, ATM, BARD1, BRIP1, CHEK2, MSH6, RAD51D, STK11*) **(Supplemental Table 1)**. A description of the genes specific to breast cancer along with their relative risk **[23]** can be found in **Table 2**. This study examines breast cancer susceptibility genes in a Guatemalan hospital case series of women referred for a tumor biopsy.

We identified 73 pathogenic variants in *ATM*, *BARD1*, *BRCA1*, *BRCA2*, *CHEK2*, *MSH6*, *PALB2*, and *TP53*, of which 45 are unique. In addition to mutations in these genes known to have high or medium penetrance in breast and ovarian cancer, we identified 9 rare pathogenic variants in the low/unknown-penetrance genes *AXIN2, FH, MLH1, MUTYH, NF1, and SDHB*. Mutations in *BRCA1* accounted for 50.7% of the non-rare pathogenic variants (37/73), followed by *BRCA2* at 20.6% (15/73), *PALB2* and *TP53* at 6.9% (5/73) each, *ATM* at 5.5% (4/73), and *BARD1* and *CHEK2* at 4.1% (3/73) each **(Figure 1A, B)**.

**Figure 1.**
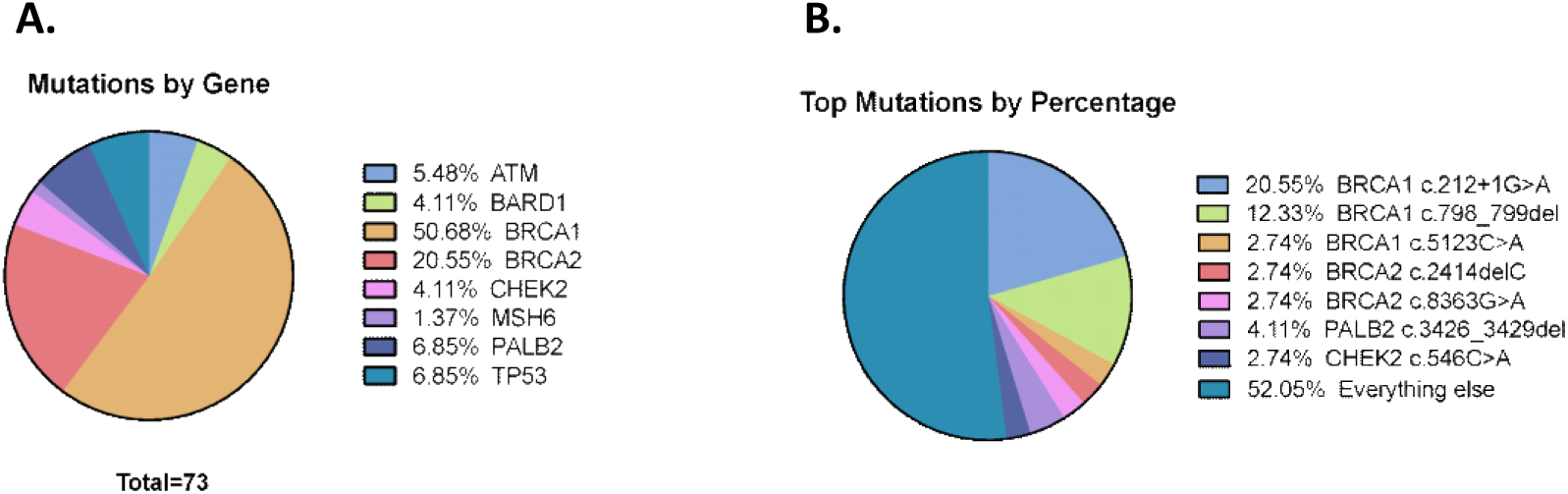
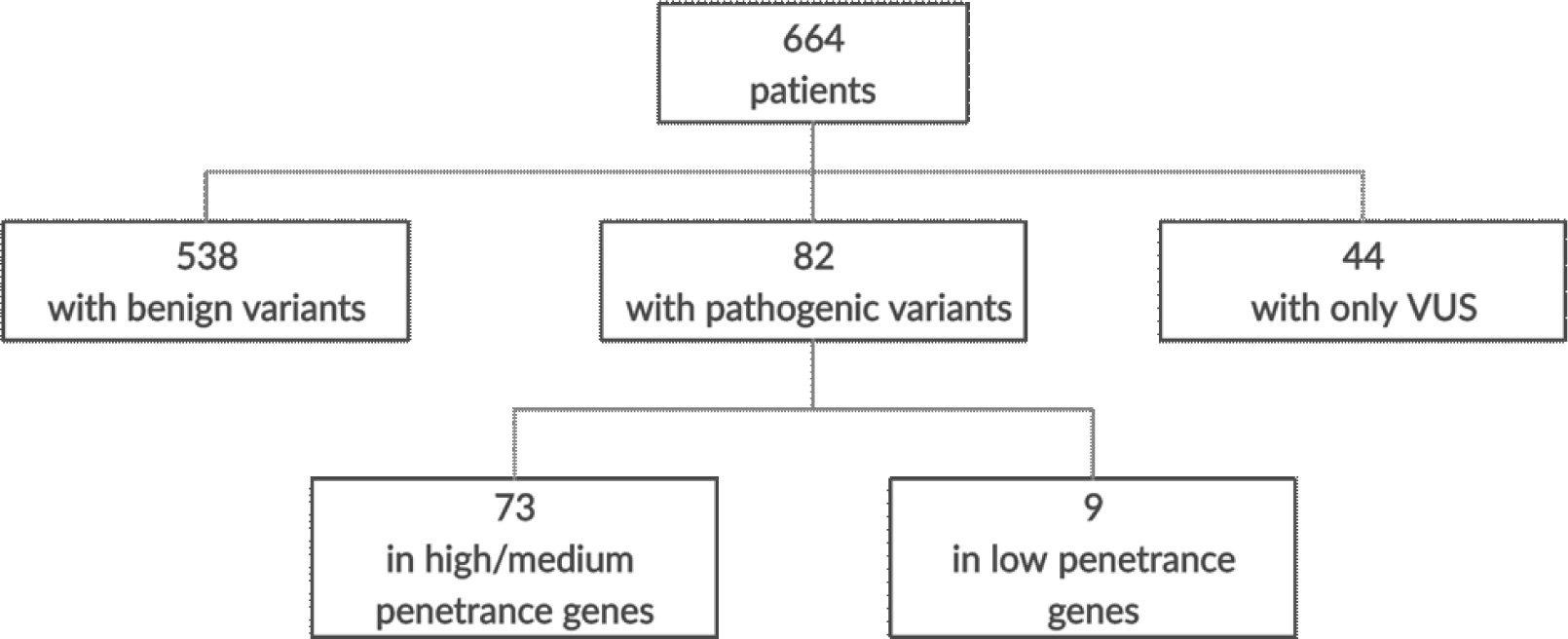
Pathogenic mutations in high and medium penetrance genes. A, Shown is the percentage of all pathogenic mutations in medium and high penetrance genes, by gene or B, by recurrent mutation. C, the groupings of patients is shown by variant type. Patient breakdown by variant type

### Association of mutations with age at diagnosis and family history

Of the 664 patients, 538 have “benign” variants (variants confirmed to be non-pathogenic), 82 have pathogenic variants, and 44 have solely variants of uncertain significance **(Figure 1C)**. Nine patients had both a pathogenic variant and variant of uncertain significance; these patients were counted in just the “pathogenic” group. The associations of study population characteristics with pathogenic mutation are observed in **Table 2**.

**Table 2.**
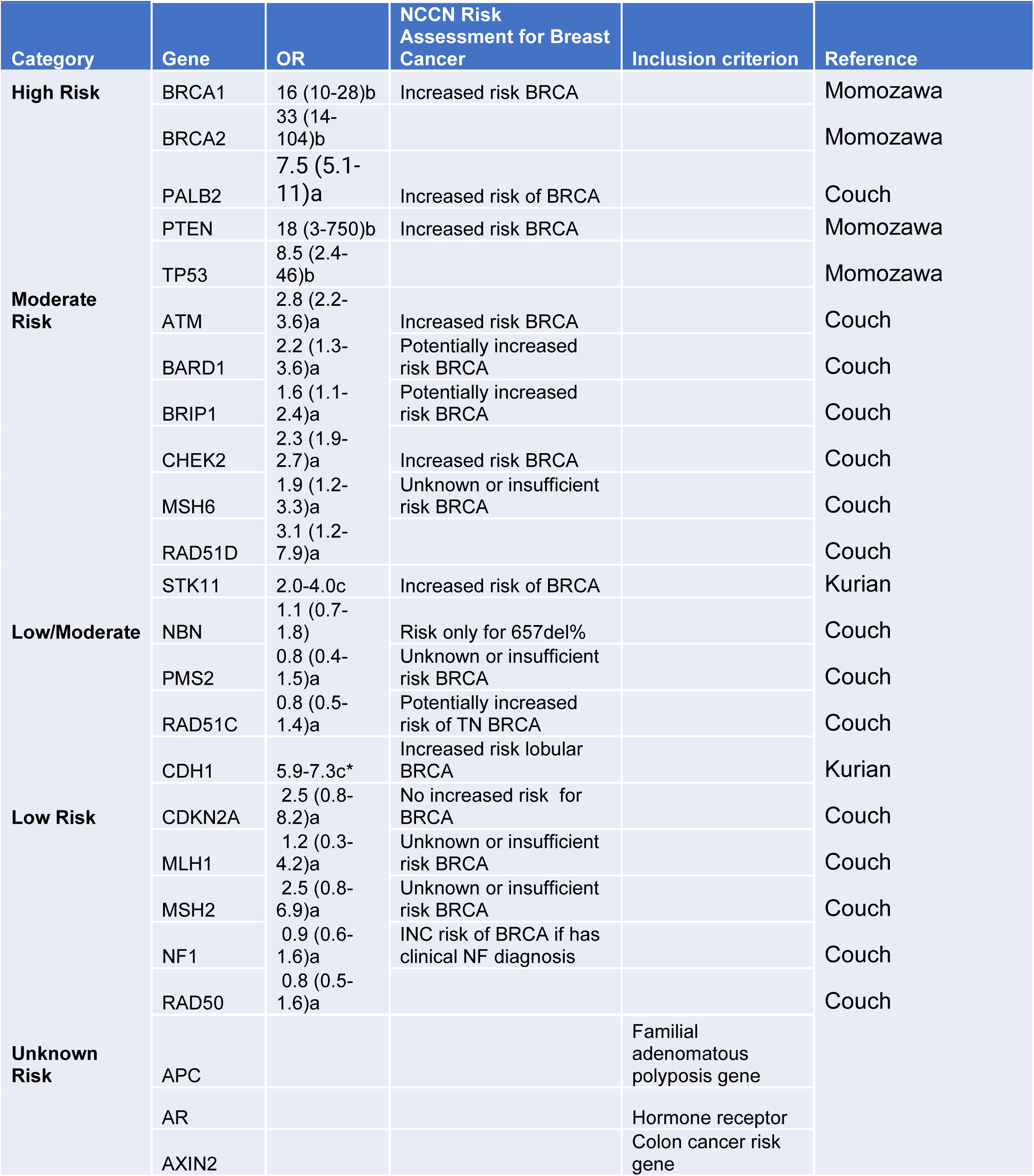
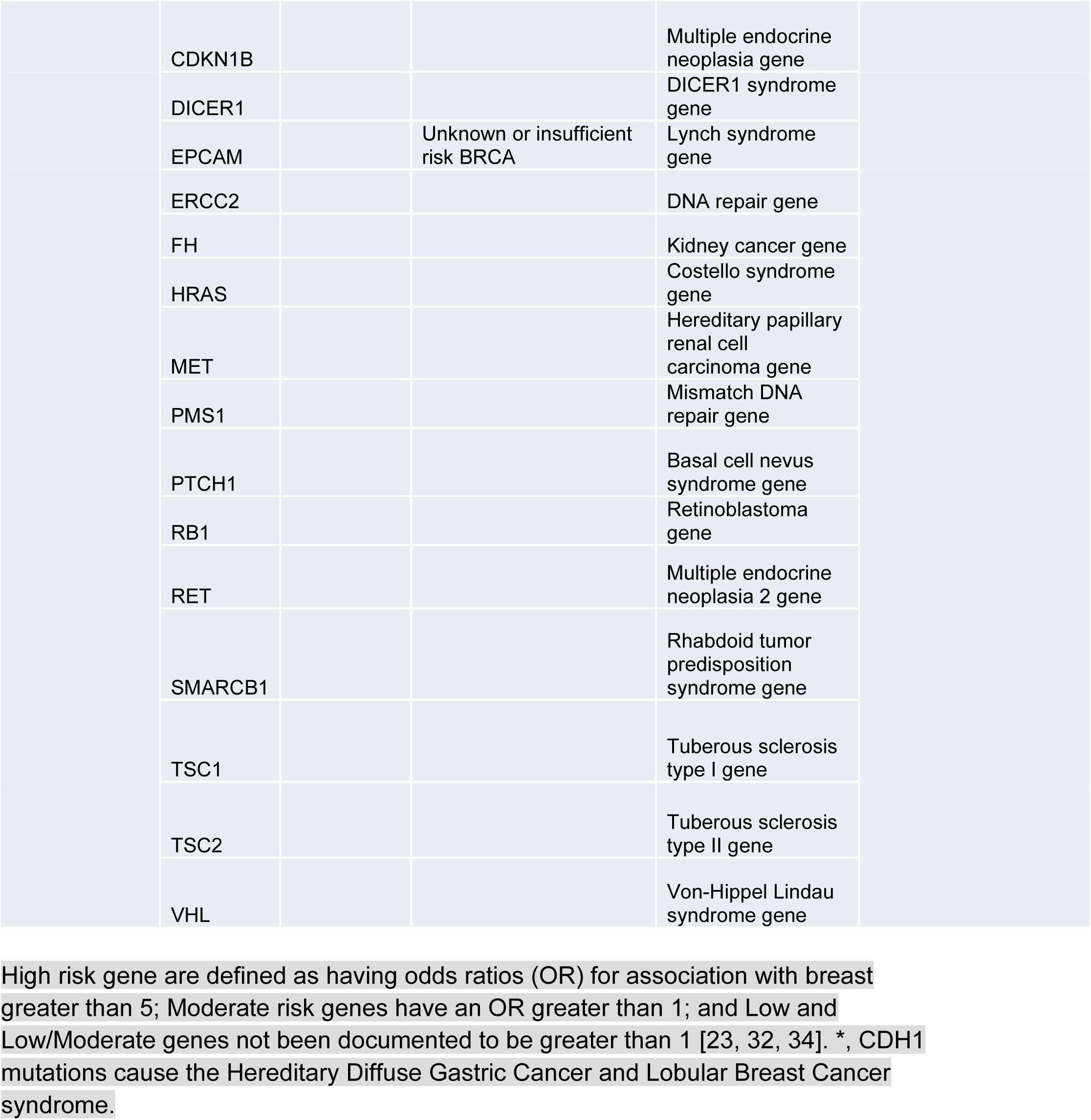
Sequenced Breast Cancer Risk Genes and their Inclusion Criteria.

Clinical characteristics of patients with pathogenic mutations in high or medium penetrance genes are compared to the remainder, that is, patients with benign variants, variants of uncertain significance, and pathogenic variants in low-penetrance genes. Patients with pathogenic mutations in high or medium penetrance genes had a statistically significant earlier age at diagnosis (median, 43 years; IQR: 36.5-51.5) compared to all other patients (median, 49 years; IQR: 41-61) (p<0.01) **(Table 1, Figure 2A)**. In addition, 33% of patients with pathogenic mutations in high or medium penetrance genes reported having a family member with breast cancer whereas just 15% of all other patients reported the same **(Figure 2B) (p<0.001)**. We found that patients with non-pathogenic mutations were more likely to report having had menopause before diagnosis than patients with pathogenic mutations and also found that patients with pathogenic mutations had fewer miscarriages **(Figure 2A)**. The complete clinical and pathological characteristics of the 73 pathogenic variants in high and medium penetrance genes are shown in **Supplemental Table 3**. Although factors such as the number of pregnancies and breastfeeding have been reported to affect breast cancer risk [24], we found no significant difference in these variables between the patients with benign mutations and patients those with pathogenic mutations.

**Figure 2.**
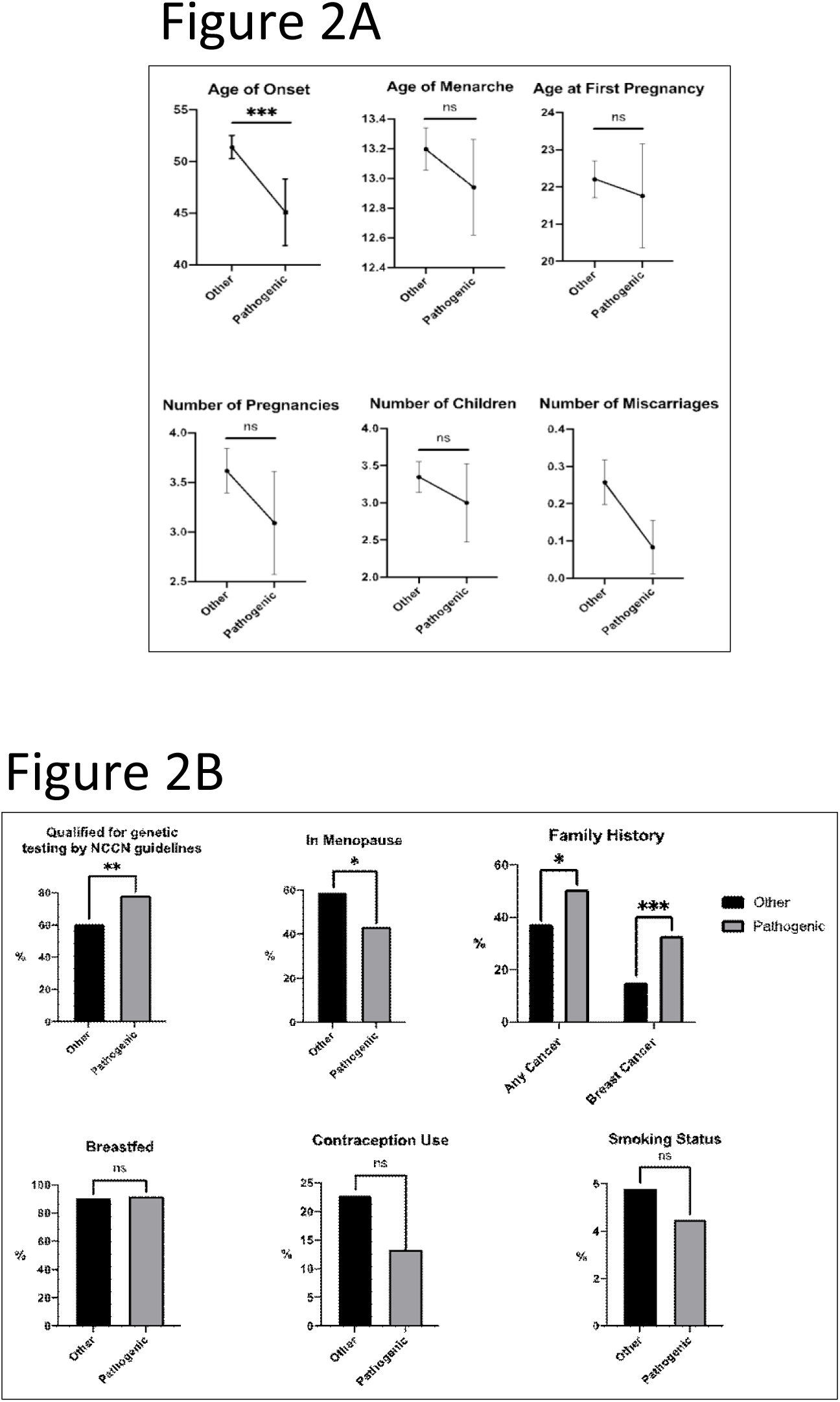
Comparisons of subjects with pathogenic mutations in high and medium penetrance genes to all other subjects. A, Subjects in the two groups alone with the interquartile range are shown for age at diagnosis, age at menarche, age at first pregnancy, number of pregnancies number of children, and number of miscarriages. B, Bar graphs of subjects meeting NCCN testing criteria, post-menopausal, with family history of breast cancer, breastfed at least one child, used oral contraceptives, or had any smoking history. *, p<0.01; **, p<0.001; ***, p<0.0001; ns, not significant.

The collective data of all women in our study suggests a significant difference in average age at diagnosis between Guatemalan and US women. The average age at diagnosis for the 664 Guatemalan women is 50.7 years whereas the average for US women is 62 [25].

## Recurrent Mutations

A recurrent mutation in this study is defined as any mutation occurring in more than one patient. Eight recurrent pathogenic mutations were discovered in this Guatemala population, including three in *BRCA1* (c.799delT, c.212+1G>A, c.5123C>A), two in *BRCA2* (c.8363G>A, c.2414delC), one in *CHEK2* (c.546C>A), one in *PALB2* (c.3426_3429del), and one in low-penetrance gene *MUTYH* (c.1218_1219dup) **(Figure 1B)**. The *BRCA1* variants c.212+1G>A (rs80358042) and c.799delT (rs80357724) occurred most frequently, accounting for 19% and 12% of all pathogenic mutations, respectively **(Figure 1B)**. The c.212+1G>A (rs80358042) is a splice site mutation that results in a truncated exon whereas c.798_799del (rs80357724) is a frameshift variant, therefore both mutations result in premature stop codons.

Given the high number of women with the *BRCA1* c.212+1G>A and c.798_799del, haplotype analysis was conducted to assess the ancestral origin of these mutations **(Figure 3A, B)**. Genome-wide SNP array data was available to determine the haplotype around a 62 Kb region in 14 of the *BRCA1* c.212+1G>A carriers and around a 56 Kb region in 9 of the *BRCA1* c.798_799del carriers. The *BRCA1* c.212+1G>A mutation is present on a haplotype common to both Hispanic and European populations so no further determination can be made. However, haplotype analysis revealed a likely European origin for the mutation *BRCA1* c.798_799del as the mutation is linked to the C allele of a SNP (rs1799950), present in all 9 carriers, that is much more common in European than in Amerindian or African populations. There was not a high enough number of patients nor informative SNPs to perform a similar analysis on the other recurrent mutations.

**Figure 3.**
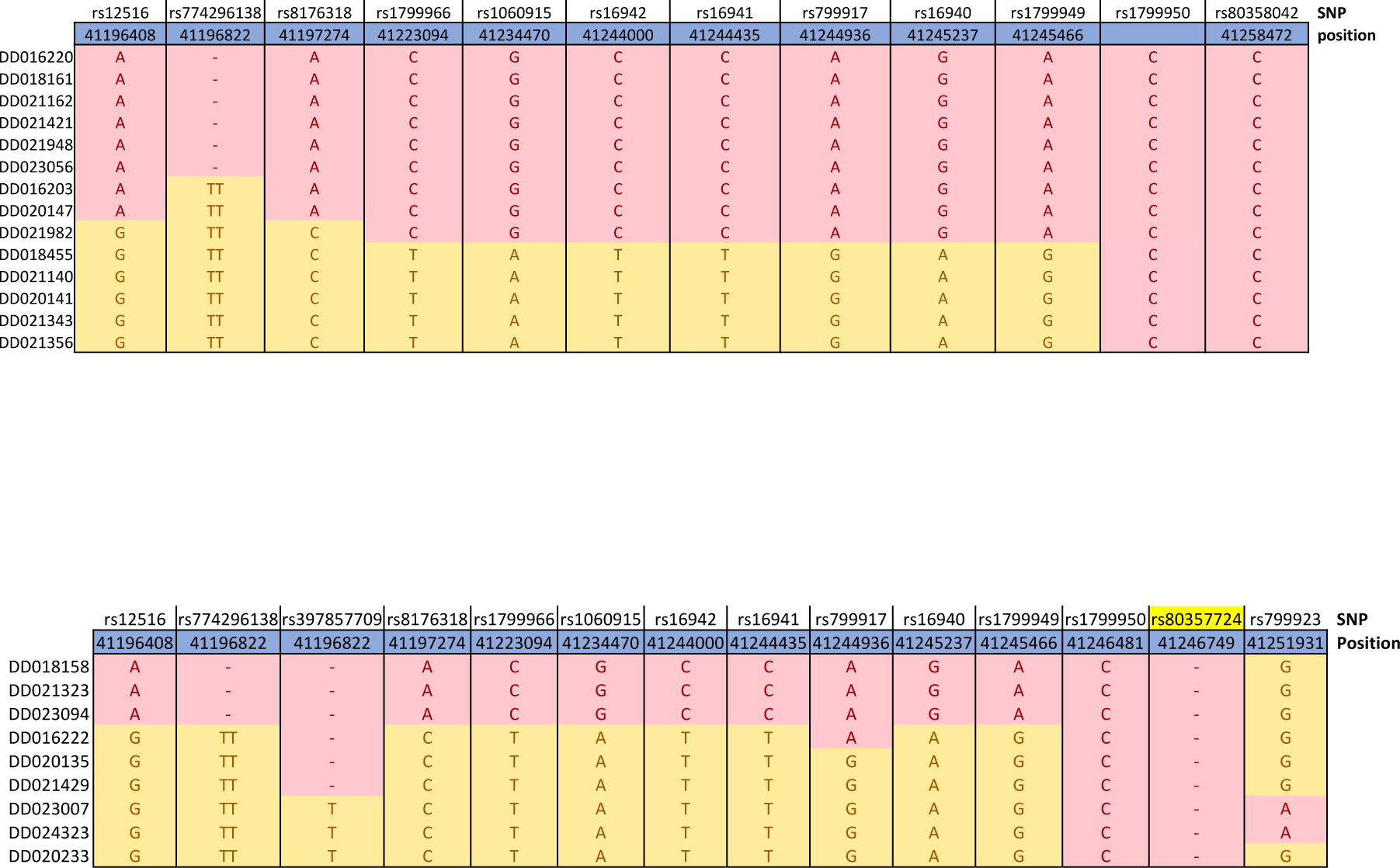
Haplotype Analysis of putative founder mutations. A, A haplotype analysis of the *BRCA1* c.212+1G>A is shown depicting each patient carrying the mutation and their genotype for linked SNPs. Phase is inferred. In red are markers that are consistent with a single haplotype present in all patients. B, The same analysis for the *BRCA1* c.798_799del mutation. Haplotype Analysis of BRCA1c.212+1G>A Haplotype Analysis of BRCA1c.798_799del

Among patients with pathogenic mutations, no significant differences in clinical presentation could be detected between those with a recurrent and a non-recurrent pathogenic mutation. However, an examination of individual mutations revealed that two of the recurrent variants have even earlier ages at diagnosis compared to the average ages at diagnosis for patients with pathogenic mutations. Thus, the average age at diagnosis for patients with *BRCA1* c.212+1G>A and *PALB2* c.3426_3429del mutations were 44 and 39 **(Supplemental Table 4)**.

### Variants of uncertain significance

Fifty-four variants were identified in high penetrance breast cancer genes that had no definitive interpretation in ClinVar or Varsome. These 54 variants of uncertain significance (VUS) were seen in 53 patients, with one patient containing 2 VUS and nine patients also carrying pathogenic variants, for a total of 44 (6.6%) unique patients with a VUS only. Thirteen of the variants were found in *BRCA1*, thirty in *BRCA2*, seven in *PALB2*, and two each in *PTEN* and *TP53*. Of these 54 VUS, 6 were recurrent mutations and 45 were unique. In ascending order, the percentage of unique variants in each high penetrance gene that is a VUS are 29% (2/7) in *TP53*, 43% (12/28) in *BRCA1*, 63% (5/8) in *PALB2*, 65% (24/37) in *BRCA2*, and 100% (2/2) in *PTEN* **(Supplemental Table 5)**.

In addition to these variants of uncertain significance, we have identified pathogenic variants in genes proposed but not demonstrated to be involved in inherited breast cancer. These genes include *AXIN2*, *FH*, *MLH1, MSH2, MUTYH, NF1*, and *SDHB*. We did not include these variants in our clinical analysis of patients with pathogenic breast cancer variants, as it is unlikely that they are directly linked to breast cancer. We also observed some rare, non-coding variants in high penetrance genes (*BRCA1, BRCA2, PALB2, TP53*). None of these variants appeared to be pathogenic from our additional risk assessment using Align-GVGD. However, it is possible that these variants may affect splicing or protein function. A list of all variants with their classifications can be found in **Supplemental Table 3**.

### Mammogram use in relation to socioeconomic status

The American Cancer Society measures breast cancer screening rate by the percentage of women 40 and older who had a mammogram in the past 2 years [14]. Of the 445 women over 40 with mammography data, 370 (83%) indicated that they have received a mammogram but only 159 (36%) have had regular screening **(Figure 4A)**. To provide more context for the rates of mammogram use, we sought to determine whether there is a connection between mammogram use and socioeconomic status. Based on a study in 2015, 80% of the Guatemalan population is estimated to work in the informal sector [26], and, therefore, we used cookstove type as a measure of SES (methods). The relation between the presence of wood-burning stoves in the house and low mammogram usage among women over 40 was extremely significant, with a p-value of 0.0004 **(Figure 4B)**. Women with wood burning stoves in their houses were less likely to have received prior mammogram screening. We also found a significant relationship between the presence of wood-burning stoves in the house and indigenous ancestry (*p* = 0.0002), indicating that women with greater genetic indigenous ancestry (ASN score of 0.5 or above) are more likely to also have a wood-burning stove at home **(Figure 4B)**.

**Figure 4.**
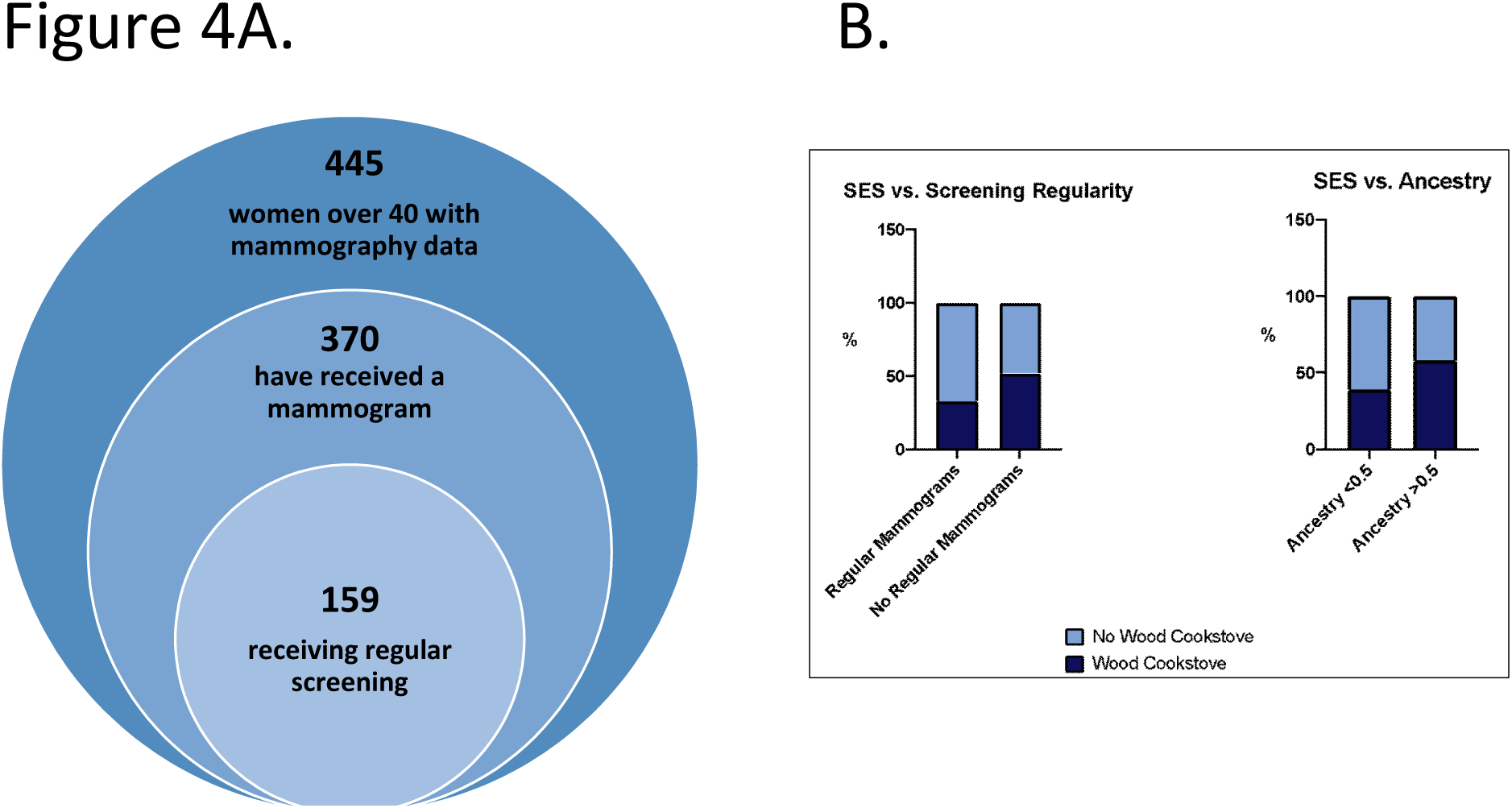
Use of mammography screening is associated with higher European ancestry and higher socioeconomic status. A, diagram showing the 445 women over 40 with mammography data, the 370 who have received screening, and the women over 40 who received (postulated) regular screening. B, bar graphs of regular mammography screening and socioeconomic status (SES), as measured by wood cookstove use (left panel) or less than or greater than 50% “Asian” ancestry (right panel).

### NCCN Guidelines for Genetic Testing

The National Comprehensive Cancer Network (NCCN) guidelines are widely recognized and used as the standard for clinical decision making. The guidelines for breast cancer screening and diagnosis take into account risk factors such as age and family history to recommend genetic testing based on each patient’s likelihood of carrying a pathogenic mutation. We thus sought to determine the utility of the NCCN guidelines in identifying Guatemalan patients with pathogenic mutations. We found that 79% of patients with pathogenic mutations in high or medium penetrance breast cancer genes met the NCCN criteria for genetic screening compared to 61% of all other patients **(Table 2, Figure 2B)**. Therefore, these guidelines will be useful for clinical genetic decision making.

## DISCUSSION

In our study of 664 breast cancer patients from Guatemala, 11% (73/664) were found to carry pathogenic mutations in high and medium penetrance breast cancer susceptibility genes. Guatemalan women have a higher ratio of deleterious *BRCA1*/*BRCA2* mutations (2.8) than US Hispanic women or other Latin American populations [3-6]. The higher *BRCA1* mutation rate is attributable to the high prevalence of the c.212+1G>A and c.799delT mutations. These two mutations are not known to be common in the United States and other Latin American countries, suggesting that they are founder mutations in the Guatemalan population [3, 6, 8]. A previous study of 222 patients across four Latin American countries (Argentina, Colombia, Guatemala, and Mexico) had described a prevalence of pathogenic variants at 17% (38/222); however, if only high and medium penetrance breast cancer genes are included that percentage drops to 13% (29/222) overall and 11% (2/19) in Guatemala [6]. The BRCA mutation prevalence among Latinas in the US is 1.2% to 4.9% in cohorts of unselected patients with breast cancer and a separate study of 1054 BRCA-negative, high-risk Hispanic women found that 4.5% carried a pathogenic variant in other cancer susceptibility genes [8, 27].

Due to the unique population history of Latin American countries and Hispanic populations, a number of recurrent mutations have been observed, including a deletion of exons 9–12 in *BRCA1* observed in Mexico and Mexican Americans, and *BRCA2* E1308X in Puerto Rico [3, 5, 7]. One recurrent mutation that has been documented in several populations across Latin America and US Hispanics is the Jewish founder mutation *BRCA1 185delAG [4, 28]*. However, this mutation was not seen in the Guatemalan population. The six recurrent mutations discovered (*BRCA1* c.212+1G>A, *BRCA1* c.799delT, *BRCA1* c.5123C>A, *BRCA2* c.8363G>A, *BRCA2* c.2414delC, and *PALB2* c.3426_3429del) seem to be uniquely recurrent within Guatemala. With increasing rates of intraregional migration, particularly to Mexico and the United States, geneticists in Central and North America should look for these mutations when screening Guatemalan patients for pathogenic variants. Particular attention should be given to *BRCA1* mutations c.212+1G>A and *BRCA1* c.799delT, which were the most prevalent within our study population. These two mutations were also seen once each in the 19 Guatemalan patients included in Oliver et al. [6].

The high percentage of variants in high and moderate penetrance genes that are classified as VUS suggest that the current online databases are still underpowered, especially in Latin American samples. As the second most commonly affected gene in hereditary breast and ovarian cancer, *BRCA2* especially warrants further study. One variant, in particular, *BRCA2* p.Leu2962_Asp2983del is notable for being an in-frame deletion of 22 amino acids. Although it is a very large deletion, it is classified as a VUS in ClinVar. We identified this deletion in 4 Guatemalan patients who do not have early-onset disease nor family history of breast cancer, suggesting this is either a benign or hypermorphic allele. Additionally, several pathogenic mutations were detected in genes with low or unknown penetrance for breast cancer. Further research is needed to clarify whether these VUS and rare pathogenic mutations are clinically actionable.

Consistent with current knowledge, patients with pathogenic mutations in known susceptibility genes had on average a lower age at diagnosis than those with benign mutations. A family history of breast cancer was also elevated in patients with pathogenic mutations. At an average of 51 years, the age of breast cancer diagnosis in our sample of Guatemalan patients was lower than the US average of 62 years [25]. Although the reason for this difference is unknown, it may suggest that women from Guatemala would benefit from starting annual mammograms at an earlier age. However, this would require a larger outreach effort to communities outside of Guatemala City and in Mayan-speaking communities.

Amerindian ancestry is associated with a lower incidence of breast cancer, mostly due to a protective allele at rs3778609 that is common in Mexican, Central, and South American populations [29]. Guatemalans have a high percentage of Amerindian ancestry and 30-40% speak one of the Mayan languages as their primary language [9]. Our study found a low percentage of women (<1%) speaking a Mayan language, lower than the 7% of women in a parallel study of cervical cancer carried out in the same clinic at INCAN [30]. A formal case-control of breast cancer in Central America is warranted to study the role of germline variants in breast cancer. Our data also has shown that Guatemalan women have significantly lower rates of breast cancer screening compared to U.S. Hispanics. In addition, women with a greater percentage of Amerindian ancestry and women of lower socioeconomic status (as indicated by cookstove type) are less likely to have previously received a mammogram. This is unsurprising as there is a great degree of overlap between these two groups. Women of higher socioeconomic status are likely to be more educated and knowledgeable about breast cancer, and therefore more likely to seek out mammograms [31]. These findings illustrate that mammogram screenings are among the health disparities to be addressed between different socioeconomic and ethnic groups.

The median age of the subjects in our study of unselected breast cancer cases is 49 years of age, considerably younger than the median age of breast cancer overall in the United States, 62 years, but similar to the mean age of 49 of women seeking genetic testing [32]. An age difference has also been observed in a study of American women undergoing testing for hereditary breast-ovarian cancer, in which women of non-European descent (mean age, 45.9 years; SD, 11.6 years) were found to be younger than European women (mean age, 50 years; SD, 11.9 years; P < .001) [4]. However, the reasons for this age difference are unknown.

From self-reported data, only 36% of Guatemalan women at these hospitals are receiving regular mammogram screening. By comparison, the breast cancer screening rate for Hispanic women in the US in 2015 was 61% [14]. This represents a 25% difference in mammogram usage and supports the conclusion that a disparity in breast cancer screening rates exists between Guatemalan women and U.S. Hispanic women. This disparity is likely explained by a less developed health care system with deficient or non-existing screening and health education programs, especially in rural areas [31].

Finally, our data show that the NCCN guidelines for breast cancer genetic testing are reasonably effective for Guatemala. A total of 79% of patients with pathogenic mutations in high or medium penetrance genes would have qualified for testing, leaving 21% unaccounted for. However, with the diversity of breast cancer patients and presentations, these guidelines may need to be adjusted on a regional or national basis.

To the best of our knowledge, this is the largest genetic study of Central American breast cancer. Our study has several limitations that should be emphasized. The population represented in this study were hospital-based case samples in the capital city and possibly skewed for women of higher socioeconomic status, women with more advanced disease, and not representative of rural populations. We in fact documented that indigenous women whose primary language is not Spanish are under-represented in our sample. This may be due to under-representation in indigenous women seeking treatment at INCAN, or in learning about and enrolling in our study [33]. Mestizo women are more likely to have received education about breast cancer and seek screening at an earlier age or perhaps at all. Another limitation of our study is that mammography history and cancer history are self-reported by patients and not verified by medical records. Finally, there is a lack of hormone receptor status and histology data that could lend insight into the specific breast cancer subtypes present in this patient population.

## CONCLUSION

A large gene panel demonstrates that *BRCA1* and *BRCA2* are the dominant breast cancer susceptibility genes in Guatemala and this data increases understanding of the molecular and demographic characteristics of breast cancer in Guatemalan women. The pathogenic variants identified, and their clinical characteristics should be used to more specifically inform cancer screening, diagnosis, and treatment of Guatemalans in the United States and abroad.

## Data Availability

DNA sequencing data will be made available to qualified researchers through dbGAP (https://www.ncbi.nlm.nih.gov/gap/).

**List of abbreviations**
IRB: institutional review board;
SES: socioeconomic status;
NCCN: National Comprehensive Cancer Network;
VUS: variant of unknown significance.

## Declarations

### Ethics approval and consent to participate

The studies wee approved by the ethics committees at INCAN and the Hospital General San Juan de Dios, all patients provided written informed consent.

### Consent for publication

Not applicable

### Authors’ contributions

AR, AA, JO, LB, SR, VA, RO, EG participated in patient recruitment and characterization, KS, BC, LW, LG, DW, CC, JW carried out sample processing, data generation and analysis, MR, CSA, performed statistical analysis, MR, MF, MD characterized genetic variants, MR and MD wrote the paper, and all authors reviewed the manuscript.

## Acknowledgements

We thank Patricia Zaid, Ester Avila, Adolfo Santizo, Lineth Boror and Luz Garcia for assistance in recruiting and consenting patients.

